# The Role of Genomic-Informed Risk Assessments in Predicting Dementia Outcomes

**DOI:** 10.1101/2024.04.27.24306488

**Authors:** Shea J. Andrews, Paulina Tolosa-Tort, Caroline Jonson, Brian Fulton-Howard, Alan E Renton, Jennifer S Yokoyama, Kristine Yaffe, the Alzheimer’s Disease Neuroimaging Initiative

**Affiliations:** Department of Psychiatry and Behavioral Sciences, University of California San Francisco, 505 Parnassus Ave, San Francisco, CA, USA, 94143; Memory and Aging Center, Department of Neurology, University of California, San Francisco, CA, USA, USA, 94143; Center for Alzheimer’s and Related Dementias, National Institutes of Health, Bethesda, MD, USA; DataTecnica LLC, Washington, DC USA; Department of Genetics and Genomic Sciences, Icahn School of Medicine at Mount Sinai, 1428 Madison Ave, New York, NY, USA, 10029; Department of Epidemiology and Biostatistics, University of California San Francisco, 505 Parnassus Ave, San Francisco, CA, USA, 94143

**Keywords:** dementia risk scores, polygenic risk scores, genomic-informed risk assessments

## Abstract

**Introduction:** By integrating genetic and clinical risk factors into genomic-informed dementia risk reports, healthcare providers can offer patients detailed risk profiles to facilitate understanding of individual risk and support the implementation of personalized strategies for promoting brain health.

**Methods:** We constructed an additive score comprising the modified Cardiovascular Risk Factors, Aging, and Incidence of Dementia Risk Score (mCAIDE), family history of dementia, *APOE* genotype, and an AD polygenic risk score in NACC and ADNI, and assessed its association with progression to all-cause dementia.

**Results:** 81% of participants had at least one high-risk indicator for dementia, with each additional risk indicator linked to a 34% increase in the hazard of dementia onset.

**Discussion:** We found that most participants in memory and aging clinics had at least one high-risk indicator for dementia. Furthermore, we observed a dose-response relationship where a greater number of risk indicators was associated with an increased risk of incident dementia.

**Research in Context:** *Systematic Review:* The authors reviewed the literature using traditional sources (e.g. PubMed), focusing on integrating genetic and non-genetic risk factors for dementia prediction. We identified a knowledge gap regarding what proportion of patients evaluated at memory and aging clinics have high-risk indicators for dementia based on their family history, monogenic mutations, *APOE* genotype, polygenic risk scores, and clinical risk factors; and what is the association of risk-indicator burden with incident dementia.

*Interpretation:* Our findings highlight that most patients possess at least one high-risk indicator for dementia, with a higher burden associated with a greater dementia risk. Compiling high-risk dementia indicators into a genomic-informed risk assessment could facilitate a better understanding of individual risk and support the implementation of personalized strategies for promoting brain health.

*Future directions:* Validation of genomic-informed risk reports is needed in longitudinal population-based studies and assessing the clinical utility of genomic risk reports to promote brain health for dementia prevention.

**Highlights:**

- Compiled Genomic-informed dementia risk report of genetic and clinical risk factors
- Most memory clinic patients have at least one high-risk indicator for dementia
- A higher risk indicator burden is associated with a greater dementia risk

## Introduction

Alzheimer’s disease (AD) is a progressive neurodegenerative disorder affecting nearly 7 million Americans [1]. This number is projected to more than double by 2060, posing an estimated economic burden exceeding one trillion dollars [1]. To date, there is no cure to stop or reverse disease progression; moreover, the recent anti-amyloid immunotherapies, which moderately delay progression, face significant barriers such as high costs and require extensive medical infrastructure for the administration and monitoring of adverse events [2]. In the absence of widely accessible and highly effective disease-modifying therapies, strategies targeting modifiable risk factors to promote brain health represent the primary approach to mitigating dementia risk [3]. Applying these strategies in a primary care setting will require risk assessment of genetic and clinical risk factors for risk stratification and communication of risk profiles for personalized risk reduction strategies [3].

Genomic-informed risk assessments are innovative and recently developed integrated profiles that compile information from clinical risk factors, family history, polygenic risk scores (PRS), and monogenic mutations [4,5]. Currently, the eMERGE consortium is evaluating the clinical utility of genomic-informed risk assessments for eleven health conditions, including asthma, diabetes, hypocholesterolemia, and obesity, within the US healthcare system [4,5]. The selection of these traits was based on PRS analytic viability, feasibility, translatability, and, critically, potential clinical actionability—a consideration previously not applicable to AD until recently. Given the demonstrated benefits of personalized multi-domain lifestyle interventions, which include improved cognitive performance and risk factor profiles among high-risk older adults recruited from a primary care setting [6], it is now timely to consider such genomic-informed dementia risk reports to promote brain health.

As AD is a complex multifactorial disease influenced by genetic and environmental factors, genomic-informed dementia risk reports should ideally integrate three components of genetic risk – family history, monogenic, and polygenic – along with clinical risk factors. The heritability of late-onset AD is estimated to be between 40-60% [7]. Highly penetrant mutations in *APP, PSEN1*, and *PSEN2* are linked to monogenic forms of AD, while the *APOE*\*ε4 allele increases the risk of sporadic AD [8]. PRS further quantify the genetic risk originating from a further 80+ AD-associated loci with small effect sizes [8]. In the absence of these specific genetic tests, a family history of AD may indicate a heightened genetic predisposition, especially for first-degree relatives of AD patients [9]. Finally, up to 14 modifiable risk factors contribute substantially – approximately 45% – to AD risk, with dementia risk scores quantifying individual risk arising from environmental factors related to clinical, lifestyle, and behavioral risk factors [10–12].

The objective of this study was to determine how established genetic – including monogenic mutations, *APOE* genotype, family history, and PRS – and clinical risk factors jointly contribute to the risk of incident dementia in non-demented patients evaluated in memory and aging clinics. Although this clinical cohort is not population⍰representative, it captures an early stage in the clinical trajectory and often marks the first specialist referral after primary care. Our findings provide a foundational step toward informing the development and validation of genomic-informed risk assessment for AD that combine early⍰life genetic architecture with routinely collected clinical data to estimate the risk of progression to dementia.

## Methods

### Cohort

We used the genetic and phenotypic data from participants who contributed to the National Alzheimer’s Coordinating Center (NACC) database and Alzheimer’s Disease Neuroimaging Initiative (ADNI). The NACC consists of over 45,000 participants from 30+ past and present US-based Alzheimer Disease Core Centers and Alzheimer Disease Research Centers funded by the National Institute on Aging [13]. ADNI was launched in 2003 as a public-private partnership with the primary goal of testing whether serial magnetic resonance imaging (MRI), positron emission tomography (PET), other biological markers, and clinical and neuropsychological assessments can be combined to measure the progression of mild cognitive impairment (MCI) and early AD [14]. Participants provided informed consent, and institutional review board approval was locally obtained.

We analyzed longitudinal data for participants with 2+ visits, who were cognitively unimpaired (CU) or had a primary diagnosis of MCI at baseline, who were at least age 55 at their initial visit or whose estimated age-of-onset of cognitive impairment was at least 55, and who had been whole genome-sequenced. Diagnostic criteria for NACC and ADNI have been previously described [13,14].

### Demographic and Clinical Risk Factors

#### Race/ethnicity

Race and ethnicity were self-reported by study participants, with categories defined by the National Institutes of Health, including American Indian or Alaska Native, Asian, Black or African American, Native Hawaiian or Other Pacific Islander, and White. Ethnicity categories included Hispanic or Latino or not Hispanic or Latino. If individuals did not identify with these racial and ethnic categories, they could report “other.”

#### Modified Cardiovascular Risk Factors, Aging, and Incidence of Dementia Risk Score

The Cardiovascular Risk Factors, Aging, and Incidence of Dementia Risk Score (CAIDE) was developed in a Finnish population-based cohort to estimate 20-year dementia risk based on an individual’s midlife risk factor profile, including age, gender, education, systolic blood pressure, body mass index, total cholesterol, and physical activity [15]. The Modified Cardiovascular Risk Factors, Aging, and Incidence of Dementia Risk Score (mCAIDE) recalibrated the CAIDE risk score to predict late-life dementia in a diverse US population using the same risk factors [16]. The mCAIDE risk scores for each participant were calculated using the published equations for mCAIDE using the following variables: age, sex, education, hypertension, obesity, and hypercholesteremia. We utilized self-reported data for age, sex, and educational attainment. Obesity was defined as a Body mass index (BMI) >30, and hypertension as a sitting systolic blood pressure >140 mmHg. For NACC, hypercholesteremia was identified through self-reported medical history or clinician assessment. In ADNI, it was determined by a fasting total cholesterol level exceeding 6.21 mmol/L. Physical activity assessments were unavailable; however, CAIDE remains predictive of dementia when physical activity is not included [17]. Across the combined cohort, missingness for hypertension, hypercholesteremia, BMI, and education were 0.12%, 5.7%, 0.29%, and 4.6%, respectively. Missing data was imputed using a Random Forrest algorithm via the ‘MissForest’ R package, which implements a non-parametric method for imputing missing values for both continuous and categorical data simultaneously within a multiple imputation framework [18]. A mCAIDE score of ≥ 6 was determined to be “high-risk”.

#### Family History of Dementia

A family history of dementia (FHx) was determined based on at least one self-reported first-degree blood relative living/lived with dementia. Participants with a family history of dementia were determined to be “high-risk”.

### Genetic Risk Factors

#### Whole-Genome Sequencing

Whole genome-sequencing data for NACC and ADNI participants was generated by the Alzheimer’s Disease Sequencing Project (ADSP). ADSP is a collaborative research effort that seeks to identify novel genetic risk factors for AD [19]. The data collected and generated through the ADSP included whole exome sequencing and whole genome sequencing (WGS) from family, case-control, and cohort study designs [19]. The Release 4 WGS dataset contains 35,569 participants from 40 cohorts that have undergone standardized data management pipelines for variant calling and quality control [20,21]. Briefly, samples were sequenced by multiple centers with different platforms and libraries. The Genome Center for Alzheimer’s Disease (GCAD) mapped short reads against the hg38 reference genome using BWA MEM, called variants using the GATK HaplotypeCaller for each sample, and subsequently jointly called genotypes across all samples using GATK. QC checks included those without a GATK pass, monomorphic across all samples, or low call rate across all studies (<80%), DP<10, GQ<20, mean average depth >500, and ABhet ratio outside of 0.25 to 0.75.

Variants flagged by GCAD for removal were excluded, with additional variant and sample QC conducted using GenoTools [22]. Variants were excluded if the call rate <0.95, not in Hardy– Weinberg equilibrium (p < 1×10-4); samples were excluded if the call rate was <0.95, discordant sex was reported based on X chromosome heterozygosity, cryptic relatedness, and either insufficient or excessive heterozygosity. Genetic ancestry was determined using the PGS Catalog Calculator, which projects samples onto principal components from known ancestral populations in the jointly called 1000 Genomes Projects and Human Diversity Project (1KG+HGDP) and uses a Random Forests classifier to assign participants to a continental genetic ancestry group [23].

#### Autosomal Dominant AD mutations

Carriers of monogenic AD variants will be identified by examining 220 variants in *PSEN1* (n = 191), *PSEN2* (n = 8), and *APP* (n = 21) previously implicated with Autosomal Dominant AD mutations (ADAD) [24]. WGS was used to match variants based on chromosome, position, reference allele, and alternate allele (mutant allele in contrast to the reference allele). Individuals with at least one alternate allele in the ADAD-linked variants were classified as ADAD variant carriers.

#### APOE genotype

The SNPs defining the *APOE* genotype (rs7412 & rs429358) were extracted from the WGS data and combined to define *APOE* genotype: ε4 carriers had either the ε2/ε4, ε3/ε4, or ε4/ε4 genotype.

#### AD Polygenic Risk Score

PRS-CSx was used to construct a cross-ancestry AD-PRS excluding the *APOE* region (variants located ± 250 kb from the *APOE* ε4 defining SNP rs429358) using ancestry-specific EUR [25], AFR [26], EAS [27], and AMR [28] summary statistics with ancestry-matched LD reference panels from 1000 Genomes using the ‘auto’ and ‘meta’ flags to automatically estimate the phi parameter of the inverse-variance weighted meta-analysis of the summary statistics in the Gibbs sampler [29]. Ancestry-normalized cross-ancestry AD-PRS were subsequently estimated in the whole of ADSP using the Polygenic Score Catalog Calculator using the score file generated by PRS-CSx [23]. The Ancestry normalization uses a two-step ancestry adjustment procedure that regresses out ancestry PCs from the raw PRS such that the mean and variance of the PRS distribution are consistent across all populations [30,31]. Principal components for the 1KG+HGDP reference were estimated using FRAPOSA, with ADSP projected into the reference PCA space and a Random Forest classifier used to determine the population to which each individual is genetically similar [23]. Risk stratification within ADSP, excluding participants from NACC and ADNI, was determined by establishing a percentile cutoff. This cutoff was defined as where the odds ratio for dementia risk in the high-risk group was significantly greater than 2 compared to the rest of the cohort (Supplementary Figure 1) [4]. The 85^th^ percentile was the cutoff used to determine a “high-risk” AD-PRS in NACC and ADNI. Due to sample overlap between ADSP and participants contributing to the AFR AD GWAS, we used PLINK to estimate pairwise genetic relatedness between participants in ADSP and ADGC and excluded ∼3,000 participants from PRS estimation and subsequent statistical analysis [32].

### Statistical Analysis

Baseline characteristics of the joint NACC and ADNI cohorts were summarized across participants progressing from CU or MCI to all-cause dementia and non-progressors as percentages for categorical variables and mean (SD) for continuous variables. Co-participant demographics for the entire NACC cohort at last assessment were stratified by patient diagnosis and relationship to patient (Supplementary Tables 1 & 2). To estimate the proportion of participants identified as high-risk for dementia in our genomic-informed risk assessment, we analyzed the prevalence of each risk indicator, including carriers of ADAD variants, *APOE*\*ε4 allele carriers, those with a family history of dementia, and individuals with high mCAIDE (≥6) and AD-PRS (≥85^th^ percentile). We summed the presence of these risk indicators weighted equally to derive a cumulative risk score for each participant for a score ranging from 0-5. Cox-proportional hazard models were used to estimate the risk of dementia progression associated with increasing risk burden, adjusting for cohort, race/ethnicity, and the Clinical Dementia Rating scale – Sum of Boxes (CDR-SB). Time-to-event was measured in months from baseline, with survival time defined as the time from baseline to the first diagnosis of dementia for incident cases. For participants who did not progress to dementia during follow-up, their survival time was treated as “right-censored.” Hazard Ratios (HR) with 95% confidence intervals (95% CI) were calculated for each group by comparing the hazard rates for individuals with one or more risk indicators to those with no risk indicators. The discriminative ability of the models was assessed using the concordance index (C-index). To account for death as a competing risk, Fine and Gray models were employed to estimate subdistribution hazard ratios. P-values were 2-sided with statistical significance set at less than 0.05. All analyses were performed using R version 4.3.3.

## Results

### The majority of individuals attending memory and aging clinics exhibit at least one high-risk indicator for dementia

A total of 3,429 older adults, with an average age of 75 years (SE = 7), were included (Table 1). This cohort was comprised of 59% females and had a racial composition of 75% non-Latinx White, 15% Black, 5.2% Latinx, 3.6% other races, and 0.4% Asian; 27% of these individuals were diagnosed with MCI (74% single-domain amnestic MCI). Throughout the study, which was conducted over an average of 71 months (SE = 46), 751 participants progressed from CU/MCI to all-cause dementia, while 518 participants died. The most common high-risk indicator identified was a family history of dementia, affecting 56% of the participants. This was followed by possession of at least one *APOE*\*ε4 allele (36%), a high mCAIDE score (34%), and high AD-PRS (11%). No participants carried an ADAD mutation. Most participants had at least one risk factor, with individuals who developed dementia having a significantly higher risk indicator burden (Table 1). Specifically, 18% of participants had no identified risk indicators, 39% had one, 32% had two, 9.8% had three, and 1% had four (Figure 1).

**Table 1:**
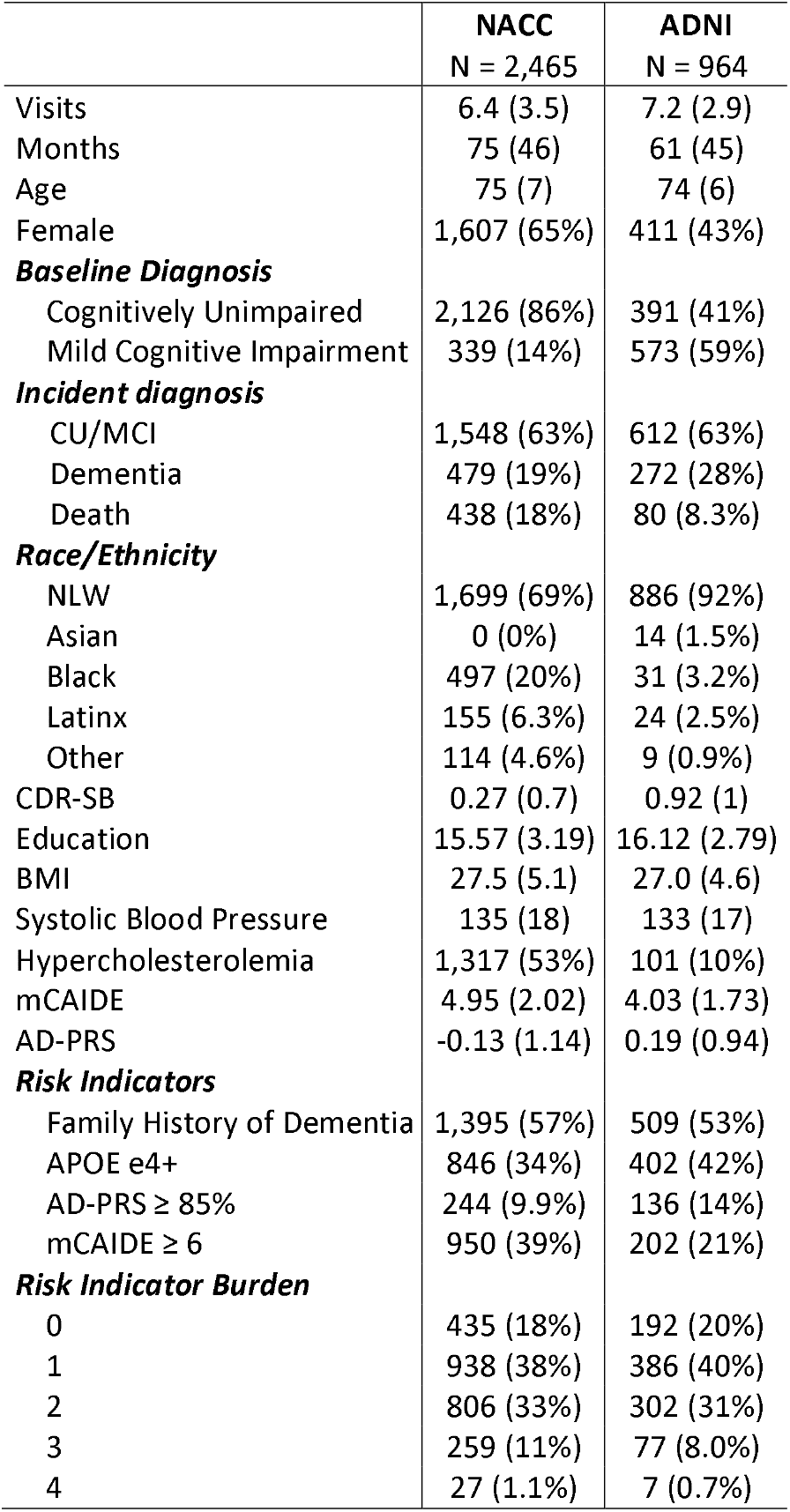
Cohort Description.

**Figure 1.**
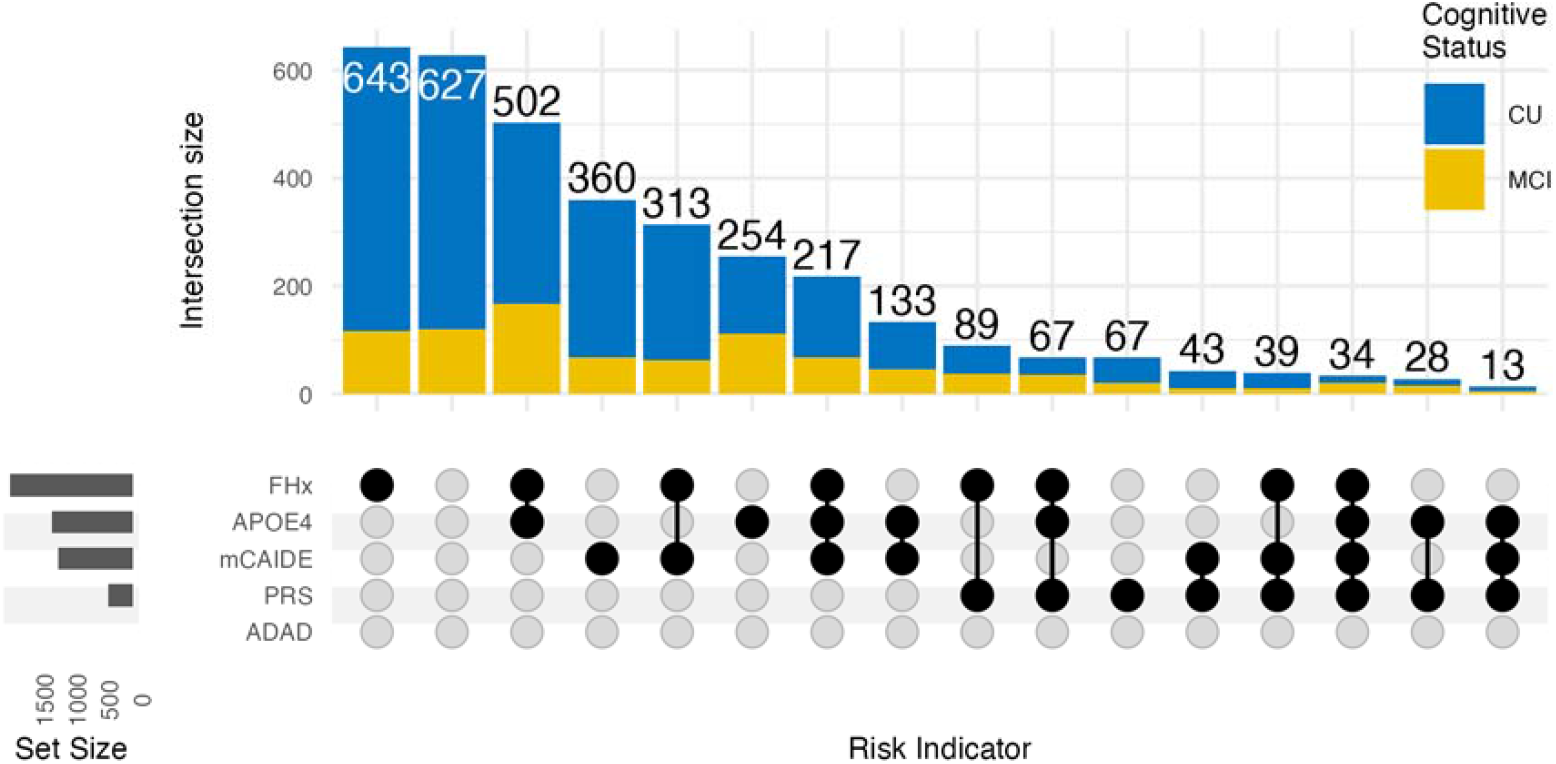
Upset plot showing the distribution and co-occurrence of dementia risk indicators at baseline among participants without dementia (i.e., the at-risk population). Horizontal bars indicate the total number of individuals with each individual risk indicator. Vertical bars represent the number of individuals with specific combinations of risk indicators. Risk indicators included: (1) family history of dementia (FHx), (2) APOE*ε4 carrier status (APOE4), (3) high (≥6) modified CAIDE risk score (mCAIDE), (4) high (top 15th percentile) Alzheimer’s disease polygenic risk score (PRS), and (5) presence of a pathogenic or likely pathogenic autosomal dominant Alzheimer’s disease (ADAD) variant in APP, PSEN1, or PSEN2. This plot reflects the baseline burden and overlap of risk components in the analytic sample prior to the development of incident dementia.

### An Increasing burden of dementia risk indicators is associated with an increased risk of dementia, exhibiting a dose-response relationship

We evaluated the association of risk indicator burden with incident dementia using Cox proportional hazard models. We observed that each additional risk indicator was linked to a 34% increase in the hazard of dementia onset (HR = 1.34, 95% CI: 1.24-1.45, p = 4e-13; c-index (SE) = 0.86 (0.006); Supplementary Table 3). Specifically, the presence of 1, 2, 3, or 4 risk indicators was associated with a 27% increase (HR = 1.27, 95% CI: 0.99-1.64, p = 0.06), 83% increase (HR = 1.83, 95% CI: 1.42-2.34, p = 2.0e-06), double (HR = 2.05, 95% CI: 1.53-2.75, p = 1.4e-06), and a fivefold (HR = 5, 95% CI: 3.02-8.29, p = 4.1e-10) greater hazard than those with no risk indicators, respectively (Figure 2; Supplementary Table 4). When accounting for death as a competing event using the Fine and Gray Model, the results remained consistent with the Cox model, though the association with a single risk indicator became statistically significant.

**Figure 2.**
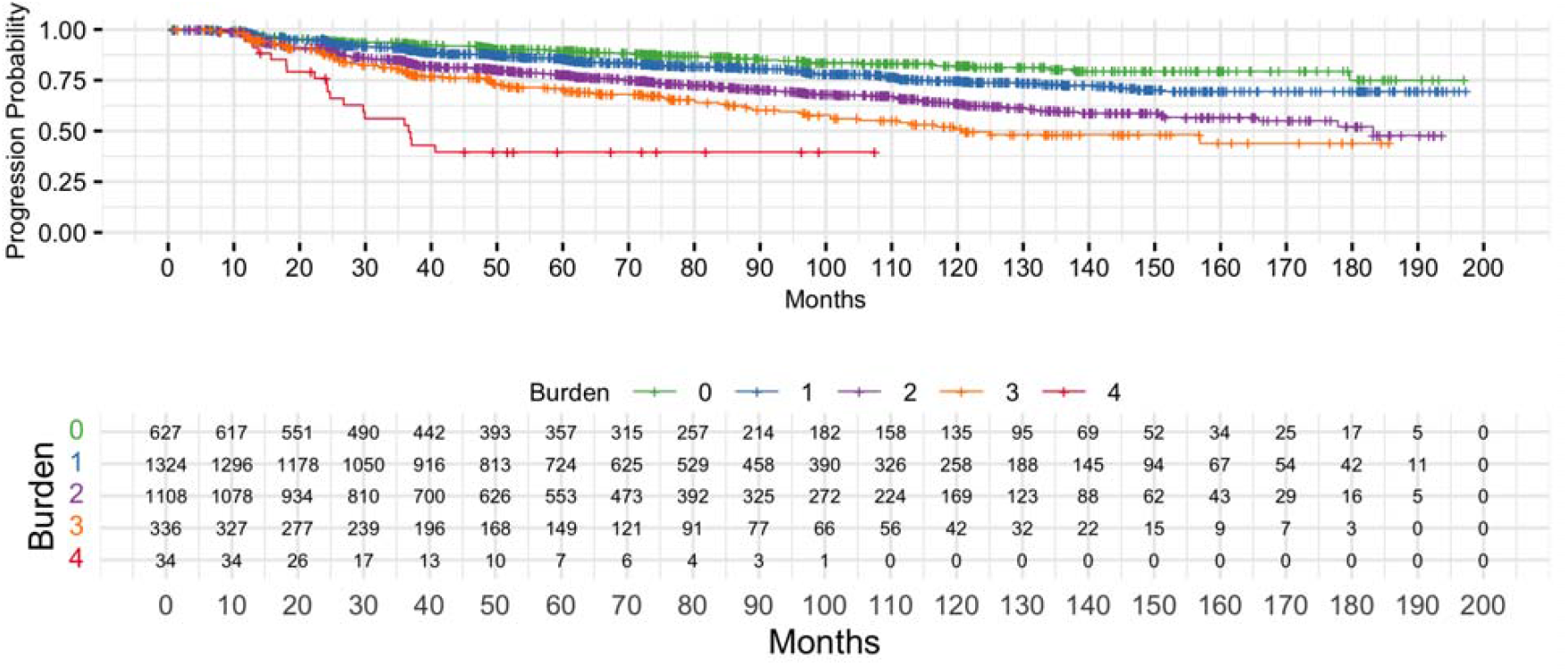
Kaplan-Meier Estimates of Time to All-Cause Dementia Onset by Number of Baseline Risk Indicators. The top panel shows Kaplan-Meier survival curves for time to progression to all-cause dementia, stratified by the number of baseline risk indicators (ranging from 0 to 4). Risk indicators included: (1) family history of dementia (FHx), (2) APOE*ε4 carrier status (APOE4), (3) high (≥6) modified CAIDE risk score (mCAIDE), (4) high (top 15th percentile) Alzheimer’s disease polygenic risk score (PRS), and (5) presence of a pathogenic or likely pathogenic autosomal dominant Alzheimer’s disease (ADAD) variant in APP, PSEN1, or PSEN2. Curves illustrate dementia-free survival over time, with higher risk burden associated with shorter time to progression to all-cause dementia. The bottom panel presents the number of individuals at risk at each follow-up interval for each risk burden group. Time is shown in months from baseline. A dose-response relationship is observed, with greater risk burden associated with faster progression to all-cause dementia.

## Discussion

Genomics⍰informed risk assessments integrate monogenic mutations, PRS, family history and clinical risk factors to give patients and healthcare providers a comprehensive view of dementia risk. In this study, we quantified the prevalence of five high⍰risk indicators—family history, ADAD mutations,⍰APOE⍰genotype, an AD⍰PRS, and mCAIDE risk score— among cognitively unimpaired or mildly impaired patients attending memory and aging clinics. Rather than designing a maximally accurate prediction model, we assessed whether the joint contribution of fixed genomic risk factors and mid-life clinical risk factors contribute to dementia progression. Although the analysis is confined to a clinical cohort, this setting mirrors the point at which most patients first meet a specialist and when preventive interventions are still feasible. We found that the majority of participants in memory and aging clinics had at least one high-risk indicator for dementia. Furthermore, we observed a dose-response relationship where a greater number of risk indicators was associated with an increased risk of incident dementia.

The ideal application for genomic-informed dementia risk assessments and personalized reports would be midlife risk assessment within primary care or family-based settings. A common trigger for interest in such an assessment is a recent parental diagnosis of dementia, which typically occurs when adult offspring are, on average, about a decade younger than the age at which blood-based biomarkers begin to show abnormality (Supplementary Tables 1 & 2; [33]). This timing creates a critical window in which genomic-informed risk assessments can identify etiology-specific vulnerability to AD/dementia, while blood-based biomarkers can later be used as stage-specific tools to detect the onset of AD pathogenesis and monitor disease progression [34]. Combining these approaches would enable precise risk stratification and clear communication of individualized risk profiles, while also highlighting modifiable factors that can be targeted to reduce overall dementia risk [3]. A staggered implementation, using genomic-informed risk assessments first, followed by a blood-based biomarker test of those at highest risk, could help optimize resource use and guide timely specialist referral. Individuals referred to dementia specialists due to abnormal blood-based biomarkers, would then undergo further evaluation using cerebrospinal fluid (CSF) and PET biomarkers [35], and, if amyloid pathology is confirmed, be considered for secondary prevention strategies, including anti-amyloid immunotherapies [36].

The clinical utility of genomic-informed risk assessments has been demonstrated for atherosclerotic cardiovascular disease (ASCVD). In a prospective cohort of middle-aged patients (n = 7,342), 42% of individuals identified as high risk for ASCVD undertook proactive measures to reduce their disease risk, subsequently improving blood lipid and blood pressure profiles [37]. This finding demonstrates that communication of personalized ASCVD risk motivates changes in health behavior and supports the integration of genomic information into clinical care for disease prevention. The ongoing GenoVA and eMERGE studies are now evaluating the clinical utility of genomic-informed risk assessments for 11 health conditions, including asthma, diabetes, hypocholesteremia, and obesity, in the U.S. healthcare system [4,5,30]. These initiatives could provide valuable insights into the development of genomic-informed dementia risk reports.

Before implementing genomic-informed risk assessments for dementia in clinical practice, several limitations need to be addressed. First, as different dementia risk scores are composed of different predictors and algorithms, the discriminative performance of different dementia risk scores needs to be validated across diverse populations. Our use of mCAIDE was determined by the lack of extensive lifestyle and social determinants of health data, precluding the use of more comprehensive dementia risk scores. Second, the predictive accuracy of PRS models is affected by the sample size ratio between the EUR and minor GWAS, between-ancestry genetic architecture differences (LD, MAF, genetic correlation, heritability, effect size), and LD reference panels [38,39]. As such, the choice of PRS model to use for portability across populations will differ between traits. We used PRS-CSx due to good overall performance when ancestry-specific GWAS from multiple populations are available; however, it is critical to assess the validity of different PRS models to determine the best approach to use [29]. Third, we treated each risk indicator as contributing to dementia risk equally in line with the eMERGE genomic-informed risk assessment design, however, more predictive models may be developed that weight each risk indicator by its relative risk. Fourth, NACC and ADNI participants — typically older, more educated, predominantly female, and with a lower prevalence of hypertension, diabetes, and depressive symptoms but a higher prevalence of subjective cognitive decline — do not represent the broader U.S. population [40,41]. This may lead to overestimates of genetic risk indicators and underestimations of environmental risk factors. As such, our findings need to be replicated in longitudinal population-based studies. Fifth, participants in this study had an average follow-up time of ∼6 years. Therefore, it is likely that many participants who developed AD already had underlying AD pathology that could have been better detected using traditional diagnostic methods, such as imaging and cognitive testing. Consequently, the utility of genomics-informed dementia risk assessments needs further evaluation in population-based cohorts more representative of the general population with longer follow-up. Finally, before genomic-informed risk assessments can be routinely applied in clinical settings, it is crucial to validate the performance of both clinical and polygenic risk scores across diverse populations. Moreover, strategies for effectively communicating these risk assessments to patients must be developed and tested to ensure they are understood and can inform patient care appropriately.

Despite these limitations, our study has several strengths. First, we have leveraged a diverse cohort with WGS and clinical evaluations, enabling us to assess ADAD mutations, *APOE* genotype, AD-PRS, a family history of dementia, and clinical risk factors. As such, we are the first study to evaluate the utility of integrated risk reports within the framework of genomic-informed risk assessments for dementia risk assessment. Second, we are using a clinical risk score, mCAIDE, validated to predict late-life dementia in a diverse U.S. population. Third, we developed a cross-ancestry AD-PRS using PRS-CSx, a state-of-the-art PRS algorithm, and the largest publicly available ancestry-specific AD GWAS summary statistics that are independent of ADSP. Additionally, the AD-PRS and the “high-risk” cut-off were developed using the full ADSP cohort, excluding NACC and ADNI, thereby enhancing the external validity of our findings.

In conclusion, most participants in memory and aging clinics possess at least one high-risk indicator for dementia. Furthermore, a higher burden of risk indicators is significantly associated with a higher likelihood of developing dementia. By integrating genetic and clinical risk factors into genomic-informed dementia risk reports, healthcare providers can offer patients detailed risk profiles. This comprehensive approach not only facilitates a better understanding of individual risk but also supports the implementation of personalized strategies for promoting brain health.

## Data Availability

All data produced in the present study are available upon reasonable request to the authors

https://naccdata.org/

https://adni.loni.usc.edu/

https://www.niagads.org/

## NACC

The NACC database is funded by NIA/NIH Grant U24 AG072122. NACC data are contributed by the NIA-funded ADRCs: P30 AG062429 (PI James Brewer, MD, PhD), P30 AG066468 (PI Oscar Lopez, MD), P30 AG062421 (PI Bradley Hyman, MD, PhD), P30 AG066509 (PI Thomas Grabowski, MD), P30 AG066514 (PI Mary Sano, PhD), P30 AG066530 (PI Helena Chui, MD), P30 AG066507 (PI Marilyn Albert, PhD), P30 AG066444 (PI John Morris, MD), P30 AG066518 (PI Jeffrey Kaye, MD), P30 AG066512 (PI Thomas Wisniewski, MD), P30 AG066462 (PI Scott Small, MD), P30 AG072979 (PI David Wolk, MD), P30 AG072972 (PI Charles DeCarli, MD), P30 AG072976 (PI Andrew Saykin, PsyD), P30 AG072975 (PI David Bennett, MD), P30 AG072978 (PI Neil Kowall, MD), P30 AG072977 (PI Robert Vassar, PhD), P30 AG066519 (PI Frank LaFerla, PhD), P30 AG062677 (PI Ronald Petersen, MD, PhD), P30 AG079280 (PI Eric Reiman, MD), P30 AG062422 (PI Gil Rabinovici, MD), P30 AG066511 (PI Allan Levey, MD, PhD), P30 AG072946 (PI Linda Van Eldik, PhD), P30 AG062715 (PI Sanjay Asthana, MD, FRCP), P30 AG072973 (PI Russell Swerdlow, MD), P30 AG066506 (PI Todd Golde, MD, PhD), P30 AG066508 (PI Stephen Strittmatter, MD, PhD), P30 AG066515 (PI Victor Henderson, MD, MS), P30 AG072947 (PI Suzanne Craft, PhD), P30 AG072931 (PI Henry Paulson, MD, PhD), P30 AG066546 (PI Sudha Seshadri, MD), P20 AG068024 (PI Erik Roberson, MD, PhD), P20 AG068053 (PI Justin Miller, PhD), P20 AG068077 (PI Gary Rosenberg, MD), P20 AG068082 (PI Angela Jefferson, PhD), P30 AG072958 (PI Heather Whitson, MD), P30 AG072959 (PI James Leverenz, MD).

## ADNI

* Data used in preparation of this article were obtained from the Alzheimer’s Disease Neuroimaging Initiative (ADNI) database (adni.loni.usc.edu). As such, the investigators within the ADNI contributed to the design and implementation of ADNI and/or provided data but did not participate in analysis or writing of this report. A complete listing of ADNI investigators can be found at: http://adni.loni.usc.edu/wp-content/uploads/how_to_apply/ADNI_Acknowledgement_List.pdf

Data collection and sharing for this project was funded by the Alzheimer’s Disease Neuroimaging Initiative (ADNI) (National Institutes of Health Grant U01 AG024904) and DOD ADNI (Department of Defense award number W81XWH-12-2-0012). ADNI is funded by the National Institute on Aging, the National Institute of Biomedical Imaging and Bioengineering, and through generous contributions from the following: AbbVie, Alzheimer’s Association; Alzheimer’s Drug Discovery Foundation; Araclon Biotech; BioClinica, Inc.; Biogen; Bristol-Myers Squibb Company; CereSpir, Inc.; Cogstate; Eisai Inc.; Elan Pharmaceuticals, Inc.; Eli Lilly and Company; EuroImmun; F. Hoffmann-La Roche Ltd and its affiliated company Genentech, Inc.; Fujirebio; GE Healthcare; IXICO Ltd.;Janssen Alzheimer Immunotherapy Research & Development, LLC.; Johnson & Johnson Pharmaceutical Research & Development LLC.; Lumosity; Lundbeck; Merck & Co., Inc.;Meso Scale Diagnostics, LLC.; NeuroRx Research; Neurotrack Technologies; Novartis Pharmaceuticals Corporation; Pfizer Inc.; Piramal Imaging; Servier; Takeda Pharmaceutical Company; and Transition Therapeutics. The Canadian Institutes of Health Research is providing funds to support ADNI clinical sites in Canada. Private sector contributions are facilitated by the Foundation for the National Institutes of Health (www.fnih.org). The grantee organization is the Northern California Institute for Research and Education, and the study is coordinated by the Alzheimer’s Therapeutic Research Institute at the University of Southern California. ADNI data are disseminated by the Laboratory for Neuro Imaging at the University of Southern California.

## ADSP

The Alzheimer’s Disease Sequencing Project (ADSP) is comprised of two Alzheimer’s Disease (AD) genetics consortia and three National Human Genome Research Institute (NHGRI) funded Large Scale Sequencing and Analysis Centers (LSAC). The two AD genetics consortia are the Alzheimer’s Disease Genetics Consortium (ADGC) funded by NIA (U01 AG032984), and the Cohorts for Heart and Aging Research in Genomic Epidemiology (CHARGE) funded by NIA (R01 AG033193), the National Heart, Lung, and Blood Institute (NHLBI), other National Institute of Health (NIH) institutes and other foreign governmental and non-governmental organizations. The Discovery Phase analysis of sequence data is supported through UF1AG047133 (to Drs. Schellenberg, Farrer, Pericak-Vance, Mayeux, and Haines); U01AG049505 to Dr. Seshadri; U01AG049506 to Dr. Boerwinkle; U01AG049507 to Dr. Wijsman; and U01AG049508 to Dr. Goate and the Discovery Extension Phase analysis is supported through U01AG052411 to Dr. Goate, U01AG052410 to Dr. Pericak-Vance and U01 AG052409 to Drs. Seshadri and Fornage.

Sequencing for the Follow Up Study (FUS) is supported through U01AG057659 (to Drs. PericakVance, Mayeux, and Vardarajan) and U01AG062943 (to Drs. Pericak-Vance and Mayeux). Data generation and harmonization in the Follow-up Phase is supported by U54AG052427 (to Drs. Schellenberg and Wang). The FUS Phase analysis of sequence data is supported through U01AG058589 (to Drs. Destefano, Boerwinkle, De Jager, Fornage, Seshadri, and Wijsman), U01AG058654 (to Drs. Haines, Bush, Farrer, Martin, and Pericak-Vance), U01AG058635 (to Dr. Goate), RF1AG058066 (to Drs. Haines, Pericak-Vance, and Scott), RF1AG057519 (to Drs. Farrer and Jun), R01AG048927 (to Dr. Farrer), and RF1AG054074 (to Drs. Pericak-Vance and Beecham).

The ADGC cohorts include: Adult Changes in Thought (ACT) (U01 AG006781, U19 AG066567), the Alzheimer’s Disease Research Centers (ADRC) (P30 AG062429, P30 AG066468, P30 AG062421, P30 AG066509, P30 AG066514, P30 AG066530, P30 AG066507, P30 AG066444, P30 AG066518, P30 AG066512, P30 AG066462, P30 AG072979, P30 AG072972, P30 AG072976, P30 AG072975, P30 AG072978, P30 AG072977, P30 AG066519, P30 AG062677, P30 AG079280, P30 AG062422, P30 AG066511, P30 AG072946, P30 AG062715, P30 AG072973, P30 AG066506, P30 AG066508, P30 AG066515, P30 AG072947, P30 AG072931, P30 AG066546, P20 AG068024, P20 AG068053, P20 AG068077, P20 AG068082, P30 AG072958, P30 AG072959), the Chicago Health and Aging Project (CHAP) (R01 AG11101, RC4 AG039085, K23 AG030944), Indiana Memory and Aging Study (IMAS) (R01 AG019771), Indianapolis Ibadan (R01 AG009956, P30 AG010133), the Memory and Aging Project (MAP) (R01 AG17917), Mayo Clinic (MAYO) (R01 AG032990, U01 AG046139, R01 NS080820, RF1 AG051504, P50 AG016574), Mayo Parkinson’s Disease controls (NS039764, NS071674, 5RC2HG005605), University of Miami (R01 AG027944, R01 AG028786, R01 AG019085, IIRG09133827, A2011048), the Multi-Institutional Research in Alzheimer’s Genetic Epidemiology Study (MIRAGE) (R01 AG09029, R01 AG025259), the National Centralized Repository for Alzheimer’s Disease and Related Dementias (NCRAD) (U24 AG021886), the National Institute on Aging Late Onset Alzheimer’s Disease Family Study (NIA-LOAD) (U24 AG056270), the Religious Orders Study (ROS) (P30 AG10161, R01 AG15819), the Texas Alzheimer’s Research and Care Consortium (TARCC) (funded by the Darrell K Royal Texas Alzheimer’s Initiative), Vanderbilt University/Case Western Reserve University (VAN/CWRU) (R01 AG019757, R01 AG021547, R01 AG027944, R01 AG028786, P01 NS026630, and Alzheimer’s Association), the Washington Heights-Inwood Columbia Aging Project (WHICAP) (RF1 AG054023), the University of Washington Families (VA Research Merit Grant, NIA: P50AG005136, R01AG041797, NINDS: R01NS069719), the Columbia University Hispanic Estudio Familiar de Influencia Genetica de Alzheimer (EFIGA) (RF1 AG015473), the University of Toronto (UT) (funded by Wellcome Trust, Medical Research Council, Canadian Institutes of Health Research), and Genetic Differences (GD) (R01 AG007584). The CHARGE cohorts are supported in part by National Heart, Lung, and Blood Institute (NHLBI) infrastructure grant HL105756 (Psaty), RC2HL102419 (Boerwinkle) and the neurology working group is supported by the National Institute on Aging (NIA) R01 grant AG033193.

The CHARGE cohorts participating in the ADSP include the following: Austrian Stroke Prevention Study (ASPS), ASPS-Family study, and the Prospective Dementia Registry-Austria (ASPS/PRODEM-Aus), the Atherosclerosis Risk in Communities (ARIC) Study, the Cardiovascular Health Study (CHS), the Erasmus Rucphen Family Study (ERF), the Framingham Heart Study (FHS), and the Rotterdam Study (RS). ASPS is funded by the Austrian Science Fond (FWF) grant number P20545-P05 and P13180 and the Medical University of Graz. The ASPS-Fam is funded by the Austrian Science Fund (FWF) project I904), the EU Joint Programme – Neurodegenerative Disease Research (JPND) in frame of the BRIDGET project (Austria, Ministry of Science) and the Medical University of Graz and the Steiermärkische Krankenanstalten Gesellschaft. PRODEM-Austria is supported by the Austrian Research Promotion agency (FFG) (Project No. 827462) and by the Austrian National Bank (Anniversary Fund, project 15435. ARIC research is carried out as a collaborative study supported by NHLBI contracts (HHSN268201100005C, HHSN268201100006C, HHSN268201100007C, HHSN268201100008C, HHSN268201100009C, HHSN268201100010C, HHSN268201100011C, and HHSN268201100012C). Neurocognitive data in ARIC is collected by U01 2U01HL096812, 2U01HL096814, 2U01HL096899, 2U01HL096902, 2U01HL096917 from the NIH (NHLBI, NINDS, NIA and NIDCD), and with previous brain MRI examinations funded by R01-HL70825 from the NHLBI. CHS research was supported by contracts HHSN268201200036C, HHSN268200800007C, N01HC55222, N01HC85079, N01HC85080, N01HC85081, N01HC85082, N01HC85083, N01HC85086, and grants U01HL080295 and U01HL130114 from the NHLBI with additional contribution from the National Institute of Neurological Disorders and Stroke (NINDS). Additional support was provided by R01AG023629, R01AG15928, and R01AG20098 from the NIA. FHS research is supported by NHLBI contracts N01-HC-25195 and HHSN268201500001I. This study was also supported by additional grants from the NIA (R01s AG054076, AG049607 and AG033040 and NINDS (R01 NS017950). The ERF study as a part of EUROSPAN (European Special Populations Research Network) was supported by European Commission FP6 STRP grant number 018947 (LSHG-CT-2006-01947) and also received funding from the European Community’s Seventh Framework Programme (FP7/2007-2013)/grant agreement HEALTH-F4-2007-201413 by the European Commission under the programme “Quality of Life and Management of the Living Resources” of 5th Framework Programme (no. QLG2-CT-2002-01254). High-throughput analysis of the ERF data was supported by a joint grant from the Netherlands Organization for Scientific Research and the Russian Foundation for Basic Research (NWO-RFBR 047.017.043). The Rotterdam Study is funded by Erasmus Medical Center and Erasmus University, Rotterdam, the Netherlands Organization for Health Research and Development (ZonMw), the Research Institute for Diseases in the Elderly (RIDE), the Ministry of Education, Culture and Science, the Ministry for Health, Welfare and Sports, the European Commission (DG XII), and the municipality of Rotterdam. Genetic data sets are also supported by the Netherlands Organization of Scientific Research NWO Investments (175.010.2005.011, 911-03-012), the Genetic Laboratory of the Department of Internal Medicine, Erasmus MC, the Research Institute for Diseases in the Elderly (014-93-015; RIDE2), and the Netherlands Genomics Initiative (NGI)/Netherlands Organization for Scientific Research (NWO) Netherlands Consortium for Healthy Aging (NCHA), project 050-060-810. All studies are grateful to their participants, faculty and staff. The content of these manuscripts is solely the responsibility of the authors and does not necessarily represent the official views of the National Institutes of Health or the U.S. Department of Health and Human Services.

The FUS cohorts include: the Alzheimer’s Disease Research Centers (ADRC) (P30 AG062429, P30 AG066468, P30 AG062421, P30 AG066509, P30 AG066514, P30 AG066530, P30 AG066507, P30 AG066444, P30 AG066518, P30 AG066512, P30 AG066462, P30 AG072979, P30 AG072972, P30 AG072976, P30 AG072975, P30 AG072978, P30 AG072977, P30 AG066519, P30 AG062677, P30 AG079280, P30 AG062422, P30 AG066511, P30 AG072946, P30 AG062715, P30 AG072973, P30 AG066506, P30 AG066508, P30 AG066515, P30 AG072947, P30 AG072931, P30 AG066546, P20 AG068024, P20 AG068053, P20 AG068077, P20 AG068082, P30 AG072958, P30 AG072959), Alzheimer’s Disease Neuroimaging Initiative (ADNI) (U19AG024904), Amish Protective Variant Study (RF1AG058066), Cache County Study (R01AG11380, R01AG031272, R01AG21136, RF1AG054052), Case Western Reserve University Brain Bank (CWRUBB) (P50AG008012), Case Western Reserve University Rapid Decline (CWRURD) (RF1AG058267, NU38CK000480), CubanAmerican Alzheimer’s Disease Initiative (CuAADI) (3U01AG052410), Estudio Familiar de Influencia Genetica en Alzheimer (EFIGA) (5R37AG015473, RF1AG015473, R56AG051876), Genetic and Environmental Risk Factors for Alzheimer Disease Among African Americans Study (GenerAAtions) (2R01AG09029, R01AG025259, 2R01AG048927), Gwangju Alzheimer and Related Dementias Study (GARD) (U01AG062602), Hillblom Aging Network (2014-A-004-NET, R01AG032289, R01AG048234), Hussman Institute for Human Genomics Brain Bank (HIHGBB) (R01AG027944, Alzheimer’s Association “Identification of Rare Variants in Alzheimer Disease”), Ibadan Study of Aging (IBADAN) (5R01AG009956), Longevity Genes Project (LGP) and LonGenity (R01AG042188, R01AG044829, R01AG046949, R01AG057909, R01AG061155, P30AG038072), Mexican Health and Aging Study (MHAS) (R01AG018016), Multi-Institutional Research in Alzheimer’s Genetic Epidemiology (MIRAGE) (2R01AG09029, R01AG025259, 2R01AG048927), Northern Manhattan Study (NOMAS) (R01NS29993), Peru Alzheimer’s Disease Initiative (PeADI) (RF1AG054074), Puerto Rican 1066 (PR1066) (Wellcome Trust (GR066133/GR080002), European Research Council (340755)), Puerto Rican Alzheimer Disease Initiative (PRADI) (RF1AG054074), Reasons for Geographic and Racial Differences in Stroke (REGARDS) (U01NS041588), Research in African American Alzheimer Disease Initiative (REAAADI) (U01AG052410), the Religious Orders Study (ROS) (P30 AG10161, P30 AG72975, R01 AG15819, R01 AG42210), the RUSH Memory and Aging Project (MAP) (R01 AG017917, R01 AG42210Stanford Extreme Phenotypes in AD (R01AG060747), University of Miami Brain Endowment Bank (MBB), University of Miami/Case Western/North Carolina A&T African American (UM/CASE/NCAT) (U01AG052410, R01AG028786), and Wisconsin Registry for Alzheimer’s Prevention (WRAP) (R01AG027161 and R01AG054047).

The four LSACs are: the Human Genome Sequencing Center at the Baylor College of Medicine (U54 HG003273), the Broad Institute Genome Center (U54HG003067), The American Genome Center at the Uniformed Services University of the Health Sciences (U01AG057659), and the Washington University Genome Institute (U54HG003079). Genotyping and sequencing for the ADSP FUS is also conducted at John P. Hussman Institute for Human Genomics (HIHG) Center for Genome Technology (CGT).

Biological samples and associated phenotypic data used in primary data analyses were stored at Study Investigators institutions, and at the National Centralized Repository for Alzheimer’s Disease and Related Dementias (NCRAD, U24AG021886) at Indiana University funded by NIA. Associated Phenotypic Data used in primary and secondary data analyses were provided by Study Investigators, the NIA funded Alzheimer’s Disease Centers (ADCs), and the National Alzheimer’s Coordinating Center (NACC, U24AG072122) and the National Institute on Aging Genetics of Alzheimer’s Disease Data Storage Site (NIAGADS, U24AG041689) at the University of Pennsylvania, funded by NIA. Harmonized phenotypes were provided by the ADSP Phenotype Harmonization Consortium (ADSP-PHC), funded by NIA (U24 AG074855, U01 AG068057 and R01 AG059716) and Ultrascale Machine Learning to Empower Discovery in Alzheimer’s Disease Biobanks (AI4AD, U01 AG068057). This research was supported in part by the Intramural Research Program of the National Institutes of health, National Library of Medicine. Contributors to the Genetic Analysis Data included Study Investigators on projects that were individually funded by NIA, and other NIH institutes, and by private U.S. organizations, or foreign governmental or nongovernmental organizations.

The ADSP Phenotype Harmonization Consortium (ADSP-PHC) is funded by NIA (U24 AG074855, U01 AG068057 and R01 AG059716). The harmonized cohorts within the ADSP-PHC include:Lrthe Anti-Amyloid Treatment in Asymptomatic Alzheimer’s study (A4 Study), a secondary prevention trial in preclinical Alzheimer’s disease, aiming to slow cognitive decline associated with brain amyloid accumulation in clinically normal older individuals. The A4 Study is funded by a public-private-philanthropic partnership, including funding from the National Institutes of Health-National Institute on Aging, Eli Lilly and Company, Alzheimer’s Association, Accelerating Medicines Partnership, GHR Foundation, an anonymous foundation and additional private donors, with in-kind support from Avid and Cogstate. The companion observational Longitudinal Evaluation of Amyloid Risk and Neurodegeneration (LEARN) Study is funded by the Alzheimer’s Association and GHR Foundation. The A4 and LEARN Studies are led by Dr. Reisa Sperling at Brigham and Women’s Hospital, Harvard Medical School and Dr. Paul Aisen at the Alzheimer’s Therapeutic Research Institute (ATRI), University of Southern California. The A4 and LEARN Studies are coordinated by ATRI at the University of Southern California, and the data are made available through the Laboratory for Neuro Imaging at the University of Southern California. The participants screening for the A4 Study provided permission to share their de-identified data in order to advance the quest to find a successful treatment for Alzheimer’s disease. We would like to acknowledge the dedication of all the participants, the site personnel, and all of the partnership team members who continue to make the A4 and LEARN Studies possible. The complete A4 Study Team list is available on: a4study.org/a4-study-team.; the Adult Changes in Thought study (ACT), U01 AG006781, U19 AG066567; Alzheimer’s Disease Neuroimaging Initiative (ADNI): Data collection and sharing for this project was funded by the Alzheimer’s Disease Neuroimaging Initiative (ADNI) (National Institutes of Health Grant U01 AG024904) and DOD ADNI (Department of Defense award number W81XWH-12-2-0012). ADNI is funded by the National Institute on Aging, the National Institute of Biomedical Imaging and Bioengineering, and through generous contributions from the following: AbbVie, Alzheimer’s Association; Alzheimer’s Drug Discovery Foundation; Araclon Biotech; BioClinica, Inc.; Biogen; Bristol-Myers Squibb Company; CereSpir, Inc.; Cogstate; Eisai Inc.; Elan Pharmaceuticals, Inc.; Eli Lilly and Company; EuroImmun; F. Hoffmann-La Roche Ltd and its affiliated company Genentech, Inc.; Fujirebio; GE Healthcare; IXICO Ltd.;Janssen Alzheimer Immunotherapy Research & Development, LLC.; Johnson & Johnson Pharmaceutical Research & Development LLC.; Lumosity; Lundbeck; Merck & Co., Inc.;Meso Scale Diagnostics, LLC.; NeuroRx Research; Neurotrack Technologies; Novartis Pharmaceuticals Corporation; Pfizer Inc.; Piramal Imaging; Servier; Takeda Pharmaceutical Company; and Transition Therapeutics. The Canadian Institutes of Health Research is providing funds to support ADNI clinical sites in Canada. Private sector contributions are facilitated by the Foundation for the National Institutes of Health (www.fnih.org). The grantee organization is the Northern California Institute for Research and Education, and the study is coordinated by the Alzheimer’s Therapeutic Research Institute at the University of Southern California. ADNI data are disseminated by the Laboratory for Neuro Imaging at the University of Southern California; Estudio Familiar de Influencia Genetica en Alzheimer (EFIGA): 5R37AG015473, RF1AG015473, R56AG051876; Memory & Aging Project at Knight Alzheimer’s Disease Research Center (MAP at Knight ADRC): The Memory and Aging Project at the Knight-ADRC (Knight-ADRC). This work was supported by the National Institutes of Health (NIH) grants R01AG064614, R01AG044546, RF1AG053303, RF1AG058501, U01AG058922 and R01AG064877 to Carlos Cruchaga. The recruitment and clinical characterization of research participants at Washington University was supported by NIH grants P30AG066444, P01AG03991, and P01AG026276. Data collection and sharing for this project was supported by NIH grants RF1AG054080, P30AG066462, R01AG064614 and U01AG052410. We thank the contributors who collected samples used in this study, as well as patients and their families, whose help and participation made this work possible. This work was supported by access to equipment made possible by the Hope Center for Neurological Disorders, the Neurogenomics and Informatics Center (NGI: https://neurogenomics.wustl.edu/) and the Departments of Neurology and Psychiatry at Washington University School of Medicine; National Alzheimer’s Coordinating Center (NACC): The NACC database is funded by NIA/NIH Grant U24 AG072122. NACC data are contributed by the NIA-funded ADRCs: P30 AG062429 (PI James Brewer, MD, PhD), P30 AG066468 (PI Oscar Lopez, MD), P30 AG062421 (PI Bradley Hyman, MD, PhD), P30 AG066509 (PI Thomas Grabowski, MD), P30 AG066514 (PI Mary Sano, PhD), P30 AG066530 (PI Helena Chui, MD), P30 AG066507 (PI Marilyn Albert, PhD), P30 AG066444 (PI John Morris, MD), P30 AG066518 (PI Jeffrey Kaye, MD), P30 AG066512 (PI Thomas Wisniewski, MD), P30 AG066462 (PI Scott Small, MD), P30 AG072979 (PI David Wolk, MD), P30 AG072972 (PI Charles DeCarli, MD), P30 AG072976 (PI Andrew Saykin, PsyD), P30 AG072975 (PI David Bennett, MD), P30 AG072978 (PI Neil Kowall, MD), P30 AG072977 (PI Robert Vassar, PhD), P30 AG066519 (PI Frank LaFerla, PhD), P30 AG062677 (PI Ronald Petersen, MD, PhD), P30 AG079280 (PI Eric Reiman, MD), P30 AG062422 (PI Gil Rabinovici, MD), P30 AG066511 (PI Allan Levey, MD, PhD), P30 AG072946 (PI Linda Van Eldik, PhD), P30 AG062715 (PI Sanjay Asthana, MD, FRCP), P30 AG072973 (PI Russell Swerdlow, MD), P30 AG066506 (PI Todd Golde, MD, PhD), P30 AG066508 (PI Stephen Strittmatter, MD, PhD), P30 AG066515 (PI Victor Henderson, MD, MS), P30 AG072947 (PI Suzanne Craft, PhD), P30 AG072931 (PI Henry Paulson, MD, PhD), P30 AG066546 (PI Sudha Seshadri, MD), P20 AG068024 (PI Erik Roberson, MD, PhD), P20 AG068053 (PI Justin Miller, PhD), P20 AG068077 (PI Gary Rosenberg, MD), P20 AG068082 (PI Angela Jefferson, PhD), P30 AG072958 (PI Heather Whitson, MD), P30 AG072959 (PI James Leverenz, MD); National Institute on Aging Alzheimer’s Disease Family Based Study (NIA-AD FBS): U24 AG056270; Religious Orders Study (ROS): P30AG10161,R01AG15819, R01AG42210; Memory and Aging Project (MAP - Rush): R01AG017917, R01AG42210; Minority Aging Research Study (MARS): R01AG22018, R01AG42210; Washington Heights/Inwood Columbia Aging Project (WHICAP): RF1 AG054023;and Wisconsin Registry for Alzheimer’s Prevention (WRAP): R01AG027161 and R01AG054047. Additional acknowledgments include the National Institute on Aging Genetics of Alzheimer’s Disease Data Storage Site (NIAGADS, U24AG041689) at the University of Pennsylvania, funded by NIA.

## NIAGADS

Data for this study were prepared, archived, and distributed by the National Institute on Aging Alzheimer’s Disease Data Storage Site (NIAGADS) at the University of Pennsylvania (U24-AG041689), funded by the National Institute on Aging.

## Author Contributions

Dr. Andrews had full access to all the data in the study and takes full responsibility for the integrity of the data and the accuracy of the data analysis. The code to support the analysis of this study is available at: https://github.com/AndrewsLabUCSF/GIDRR

Concept and design: SJA

Acquisition, analysis, or interpretation of data: SJA, CJ, BFH, AER

Drafting of the Manuscript: SJA, PTT

Critical review of the manuscript for important intellectual content: PTT, KY, JSY, CJ, BFJ, AER

Statistical analysis: SJA, CJ

## Consent Statement

Participants provided informed consent, and institutional review board approval was locally obtained.

## References

[1] 2023 Alzheimer’s disease facts and figures. Alzheimer’s Dement 2023;19:1598–695. 10.1002/alz.13016.

[2] Lancet T. Lecanemab for Alzheimer’s disease: tempering hype and hope. Lancet 2022;400:1899. 10.1016/s0140-6736(22)02480-1.

[3] Frisoni GB, Altomare D, Ribaldi F, Villain N, Brayne C, Mukadam N, et al. Dementia prevention in memory clinics: recommendations from the European task force for brain health services. Lancet Regional Heal - Europe 2023:100576. 10.1016/j.lanepe.2022.100576.

[4] Linder JE, Allworth A, Bland ST, Caraballo PJ, Chisholm RL, Clayton EW, et al. Returning integrated genomic risk and clinical recommendations: The eMERGE study. Genet Med 2023;25:100006. 10.1016/j.gim.2023.100006.

[5] Lennon NJ, Kottyan LC, Kachulis C, Abul-Husn NS, Arias J, Belbin G, et al. Selection, optimization and validation of ten chronic disease polygenic risk scores for clinical implementation in diverse US populations. Nat Med 2024:1–8. 10.1038/s41591-024-02796-z.

[6] Yaffe K, Vittinghoff E, Dublin S, Peltz CB, Fleckenstein LE, Rosenberg DE, et al. Effect of Personalized Risk-Reduction Strategies on Cognition and Dementia Risk Profile Among Older Adults. JAMA Intern Med 2024;184:54–62. 10.1001/jamainternmed.2023.6279.

[7] Liu S, Bush WS, Akinyemi RO, Byrd GS, Caban-Holt AM, Rajabli F, et al. Alzheimer disease is (sometimes) highly heritable: Drivers of variation in heritability estimates for binary traits, a systematic review. medRxiv 2025:2025.04.29.25326648. 10.1101/2025.04.29.25326648.

[8] Andrews SJ, Renton AE, Fulton-Howard B, Podlesny-Drabiniok A, Marcora E, Goate AM. The complex genetic architecture of Alzheimer’s disease: novel insights and future directions. eBioMedicine 2023;90:104511. 10.1016/j.ebiom.2023.104511.

[9] Cannon-Albright LA, Foster NL, Schliep K, Farnham JM, Teerlink CC, Kaddas H, et al. Relative risk for Alzheimer disease based on complete family history. Neurology 2019;92:10.1212/WNL.0000000000007231. 10.1212/wnl.0000000000007231.

[10] Peters R, Booth A, Rockwood K, Peters J, D’Este C, Anstey KJ. Combining modifiable risk factors and risk of dementia: a systematic review and meta-analysis. Bmj Open 2019;9:bmjopen-2018-022846. 10.1136/bmjopen-2018-022846.

[11] Barnes DE, Yaffe K. The projected effect of risk factor reduction on Alzheimer’s disease prevalence. Lancet Neurol 2011;10:819–-828.

[12] Livingston G, Huntley J, Liu KY, Costafreda SG, Selbæk G, Alladi S, et al. Dementia prevention, intervention, and care: 2024 report of the Lancet standing Commission. Lancet 2024. 10.1016/s0140-6736(24)01296-0.

[13] Beekly DL, Ramos EM, Lee WW, Deitrich WD, Jacka ME, Wu J, et al. The National Alzheimer’s Coordinating Center (NACC) Database: The Uniform Data Set. Alzheimer Dis Assoc Disord 2007;21:249–58. 10.1097/wad.0b013e318142774e.

[14] Petersen RC, Aisen PS, Beckett LA, Donohue MC, Gamst AC, Harvey DJ, et al. Alzheimer’s Disease Neuroimaging Initiative (ADNI) Clinical characterization. Neurology 2010;74:201–9. 10.1212/wnl.0b013e3181cb3e25.

[15] Kivipelto M, Ngandu T, Laatikainen T, Winblad B, Soininen H, Tuomilehto J. Risk score for the prediction of dementia risk in 20 years among middle aged people: a longitudinal, population-based study. Lancet Neurology 2006;5:735–41. 10.1016/s1474-4422(06)70537-3.

[16] Tolea MI, Heo J, Chrisphonte S, Galvin JE. A Modified CAIDE Risk Score as a Screening Tool for Cognitive Impairment in Older Adults. J Alzheimer’s Dis 2021;82:1755–68. 10.3233/jad-210269.

[17] Exalto LG, Quesenberry CP, Barnes D, Kivipelto M, Biessels GJ, Whitmer RA. Midlife risk score for the prediction of dementia four decades later. Alzheimer’s Dementia J Alzheimer’s Assoc 2013;10:562–70. 10.1016/j.jalz.2013.05.1772.

[18] Stekhoven DJ, Bühlmann P. MissForest—non-parametric missing value imputation for mixed-type data. Bioinformatics 2012;28:112–8. 10.1093/bioinformatics/btr597.

[19] Beecham GW, Bis JC, Martin ER, Choi S-H, DeStefano AL, Duijn CM van, et al. The Alzheimer’s Disease Sequencing Project: Study design and sample selection. Neurology Genetics 2017;3:e194. 10.1212/nxg.0000000000000194.

[20] Naj AC, Lin H, Vardarajan BN, White S, Lancour D, Ma Y, et al. Quality control and integration of genotypes from two calling pipelines for whole genome sequence data in the Alzheimer’s disease sequencing project. Genomics 2019;111:808–18. 10.1016/j.ygeno.2018.05.004.

[21] Leung YY, Valladares O, Chou Y-F, Lin H-J, Kuzma AB, Cantwell L, et al. VCPA: genomic variant calling pipeline and data management tool for Alzheimer’s Disease Sequencing Project. Bioinformatics 2018;35:1768–70. 10.1093/bioinformatics/bty894.

[22] Vitale D, Koretsky M, Kuznetsov N, Hong S, Martin J, James M, et al. GenoTools: An Open-Source Python Package for Efficient Genotype Data Quality Control and Analysis. bioRxiv 2024:2024.03.26.586362. 10.1101/2024.03.26.586362.

[23] Lambert SA, Gil L, Jupp S, Ritchie SC, Xu Y, Buniello A, et al. The Polygenic Score Catalog as an open database for reproducibility and systematic evaluation. Nat Genet 2021;53:420–5. 10.1038/s41588-021-00783-5.

[24] Wang D, Scalici A, Wang Y, Lin H, Pitsillides A, Heard-Costa N, et al. Frequency of Variants in Mendelian Alzheimer’s Disease Genes within the Alzheimer’s Disease Sequencing Project (ADSP). medRxiv 2024:2023.10.24.23297227. 10.1101/2023.10.24.23297227.

[25] Bellenguez C, Küçükali F, Jansen IE, Kleineidam L, Moreno-Grau S, Amin N, et al. New insights into the genetic etiology of Alzheimer’s disease and related dementias. Nat Genet 2022;54:412–36. 10.1038/s41588-022-01024-z.

[26] Kunkle BW, Schmidt M, Klein H-U, Naj AC, Hamilton-Nelson KL, Larson EB, et al. Novel Alzheimer Disease Risk Loci and Pathways in African American Individuals Using the African Genome Resources Panel. JAMA Neurol 2021;78:102–13. 10.1001/jamaneurol.2020.3536.

[27] Shigemizu D, Mitsumori R, Akiyama S, Miyashita A, Morizono T, Higaki S, et al. Ethnic and trans-ethnic genome-wide association studies identify new loci influencing Japanese Alzheimer’s disease risk. Transl Psychiat 2021;11:151. 10.1038/s41398-021-01272-3.

[28] Lake J, Solsberg CW, Kim JJ, Acosta-Uribe J, Makarious MB, Li Z, et al. Multi-ancestry meta-analysis and fine-mapping in Alzheimer’s disease. Mol Psychiatry 2023:1–12. 10.1038/s41380-023-02089-w.

[29] Wang Y, Kanai M, Tan T, Kamariza M, Tsuo K, Yuan K, et al. Polygenic prediction across populations is influenced by ancestry, genetic architecture, and methodology. Cell Genom 2023;3:100408. 10.1016/j.xgen.2023.100408.

[30] Hao L, Kraft P, Berriz GF, Hynes ED, Koch C, Kumar PKV, et al. Development of a clinical polygenic risk score assay and reporting workflow. Nat Med 2022;28:1006–13. 10.1038/s41591-022-01767-6.

[31] Ge T, Irvin MR, Patki A, Srinivasasainagendra V, Lin Y-F, Tiwari HK, et al. Development and validation of a trans-ancestry polygenic risk score for type 2 diabetes in diverse populations. Genome Med 2022;14:70. 10.1186/s13073-022-01074-2.

[32] Ellis CA, Oliver KL, Harris RV, Ottman R, Scheffer IE, Mefford HC, et al. Inflation of polygenic risk scores caused by sample overlap and relatedness: Examples of a major risk of bias. Am J Hum Genet 2024;111:1805–9. 10.1016/j.ajhg.2024.07.014.

[33] Milà□Alomà M, Tosun D, Schindler SE, Hausle I, Petersen KK, Li Y, et al. Timing of Changes in Alzheimer’s Disease Plasma Biomarkers as Assessed by Amyloid and Tau PET Clocks. Ann Neurol 2025. 10.1002/ana.27285.

[34] Menendez□Gonzalez M. Implementing a tridimensional diagnostic framework for personalized medicine in neurodegenerative diseases. Alzheimer’s Dement 2025;21:e14591. 10.1002/alz.14591.

[35] Frisoni GB, Festari C, Massa F, Ramusino MC, Orini S, Aarsland D, et al. European intersocietal recommendations for the biomarker-based diagnosis of neurocognitive disorders. Lancet Neurol 2024;23:302–12. 10.1016/s1474-4422(23)00447-7.

[36] Cummings J, Apostolova L, Rabinovici GD, Atri A, Aisen P, Greenberg S, et al. Lecanemab: Appropriate Use Recommendations. J Prev Alzheimer’s Dis 2023;10:362–77. 10.14283/jpad.2023.30.

[37] Widén E, Junna N, Ruotsalainen S, Surakka I, Mars N, Ripatti P, et al. How Communicating Polygenic and Clinical Risk for Atherosclerotic Cardiovascular Disease Impacts Health Behavior: an Observational Follow-up Study. Circulation Genom Precis Medicine 2022;15:e003459. 10.1161/circgen.121.003459.

[38] Hoggart C, Choi SW, García-González J, Souaiaia T, Preuss M, O’Reilly P. BridgePRS: A powerful trans-ancestry Polygenic Risk Score method. bioRxiv 2023:2023.02.17.528938. 10.1101/2023.02.17.528938.

[39] Wang Y, Kanai M, Tan T, Kamariza M, Tsuo K, Yuan K, et al. Polygenic prediction across populations is influenced by ancestry, genetic architecture, and methodology. bioRxiv 2023:2022.12.29.522270. 10.1101/2022.12.29.522270.

[40] Rentería MA, Mobley TM, Evangelista ND, Medina LD, Deters KD, Fox□Fuller JT, et al. Representativeness of samples enrolled in Alzheimer’s disease research centers. Alzheimer’s Dement: Diagn, Assess Dis Monit 2023;15:e12450. 10.1002/dad2.12450.

[41] Gianattasio KZ, Bennett EE, Wei J, Mehrotra ML, Mosley T, Gottesman RF, et al. Generalizability of findings from a clinical sample to a community □ based sample: A comparison of ADNI and ARIC. Alzheimer’s Dementia 2021;17:1265–76. 10.1002/alz.12293.

