## Supplementary Tables and Figures for "The Role of Genomic-Informed Risk Assessments in Predicting Dementia Outcomes"

**Supplementary Table 1: NACC co-participant demographics stratified by patient diagnosis.**

|  | **CU**  N = 14,354 | | | | **MCI**  N = 5,647 | | | | **Dementia**  N = 17,281 | | | |
| --- | --- | --- | --- | --- | --- | --- | --- | --- | --- | --- | --- | --- |
|  | **Partner** | **Child** | **Sibling** | **Other** | **Partner** | **Child** | **Sibling** | **Other** | **Partner** | **Child** | **Sibling** | **Other** |
| **n** | 7,545 | 3,420 | 951 | 2,438 | 3,302 | 1,374 | 233 | 738 | 11,053 | 4,922 | 538 | 768 |
| **Patient Age^†^** | 70 (11) | 78 (10) | 70 (11) | 74 (11) | 71 (10) | 77 (10) | 71 (10) | 74 (10) | 68 (10) | 76 (10) | 64 (12) | 74 (12) |
| **Age^†^** | 70 (11) | 50 (11) | 68 (11) | 69 (13) | 69 (11) | 50 (11) | 68 (11) | 67 (13) | 66 (11) | 48 (11) | 62 (12) | 59 (16) |
| **Race** |  |  |  |  |  |  |  |  |  |  |  |  |
| NHW | 1,714 (79%) | 667 (63%) | 165 (53%) | 626 (64%) | 1,734 (82%) | 428 (59%) | 64 (50%) | 233 (54%) | 7,512 (87%) | 2,500 (64%) | 295 (66%) | 370 (65%) |
| Black | 233 (11%) | 223 (21%) | 95 (30%) | 231 (24%) | 164 (7.8%) | 172 (24%) | 47 (36%) | 144 (33%) | 468 (5.4%) | 738 (19%) | 90 (20%) | 102 (18%) |
| Hispanic | 96 (4.4%) | 84 (7.9%) | 22 (7.1%) | 54 (5.5%) | 94 (4.5%) | 79 (11%) | 8 (6.2%) | 25 (5.8%) | 306 (3.5%) | 354 (9.0%) | 26 (5.8%) | 33 (5.8%) |
| Other | 139 (6.4%) | 88 (8.3%) | 30 (9.6%) | 66 (6.8%) | 114 (5.4%) | 49 (6.7%) | 10 (7.8%) | 31 (7.2%) | 343 (4.0%) | 329 (8.4%) | 37 (8.3%) | 62 (11%) |
| Uknown | 5,363 | 2,358 | 639 | 1,461 | 1,196 | 646 | 104 | 305 | 2,424 | 1,001 | 90 | 201 |
| **Education** | 15.68 (2.95) | 15.75 (2.58) | 14.68 (3.04) | 15.54 (2.78) | 15.81 (2.79) | 15.81 (2.59) | 14.98 (2.88) | 15.22 (2.87) | 15.33 (2.86) | 15.55 (2.65) | 14.89 (2.74) | 14.96 (3.26) |
| Unknown | 5,377 | 2,395 | 640 | 1,469 | 1,211 | 664 | 102 | 302 | 2,570 | 1,101 | 101 | 206 |
| **Female** | 3,674 (49%) | 2,352 (69%) | 784 (82%) | 1,968 (81%) | 2,156 (65%) | 958 (70%) | 188 (81%) | 604 (82%) | 7,008 (63%) | 3,670 (75%) | 411 (76%) | 637 (83%) |

**Other:** Other relative; Friend, neighbor, or someone known through family, friends, work, or community; Paid caregiver, health care provider, or clinician; Other

**†** Patient and co-participant age at visit when patients were first diagnosed with MCI or dementia or at last visit if cognitively unimpaired, or for patients with a baseline diagnosis of MCI or dementia (n = 18,820), their estimated age of when cognitive decline began.

**Supplementary Table 2: NACC co-participant demographics stratified by patient diagnosis, excluding baseline MCI/dementia diagnosis.**

|  | **CU**  N = 10,551 | | | | **MCI**  N = 2,462 | | | | **Dementia**  N = 3,996 | | | |
| --- | --- | --- | --- | --- | --- | --- | --- | --- | --- | --- | --- | --- |
|  | **Partner** | **Child** | **Sibling** | **Other** | **Partner** | **Child** | **Sibling** | **Other** | **Partner** | **Child** | **Sibling** | **Other** |
| **n** | 5,480 | 2,649 | 696 | 1,762 | 1,221 | 768 | 112 | 361 | 2,420 | 1,191 | 113 | 272 |
| **Patient Age^†^** | 72 (10) | 79 (9) | 72 (10) | 76 (10) | 76 (9) | 81 (9) | 76 (9) | 79 (9) | 75 (9) | 84 (8) | 74 (12) | 81 (11) |
| **Age^†^** | 72 (11) | 51 (11) | 70 (10) | 70 (13) | 73 (10) | 53 (10) | 72 (9) | 70 (13) | 73 (9) | 55 (9) | 71 (10) | 67 (15) |
| **Race** |  |  |  |  |  |  |  |  |  |  |  |  |
| NHW | 107 (73%) | 220 (71%) | 36 (53%) | 192 (69%) | 44 (85%) | 85 (62%) | 6 (55%) | 39 (57%) | 41 (76%) | 185 (77%) | 16 (64%) | 51 (65%) |
| Black | 17 (12%) | 53 (17%) | 22 (32%) | 59 (21%) | 2 (3.8%) | 32 (23%) | 4 (36%) | 14 (20%) | 3 (5.6%) | 35 (15%) | 5 (20%) | 14 (18%) |
| Hispanic | 6 (4.1%) | 19 (6.2%) | 2 (2.9%) | 11 (3.9%) | 4 (7.7%) | 14 (10%) | 0 (0%) | 6 (8.7%) | 3 (5.6%) | 12 (5.0%) | 1 (4.0%) | 4 (5.1%) |
| Other | 17 (12%) | 16 (5.2%) | 8 (12%) | 17 (6.1%) | 2 (3.8%) | 7 (5.1%) | 1 (9.1%) | 10 (14%) | 7 (13%) | 9 (3.7%) | 3 (12%) | 9 (12%) |
| Uknown | 5,333 | 2,341 | 628 | 1,447 | 1,169 | 630 | 101 | 292 | 2,366 | 950 | 88 | 194 |
| **Education** | 15.24 (3.12) | 15.95 (2.37) | 14.51 (3.17) | 15.44 (2.73) | 15.50 (2.91) | 15.80 (2.55) | 15.58 (2.81) | 14.87 (2.76) | 15.24 (3.04) | 16.16 (2.34) | 14.56 (2.14) | 15.09 (2.59) |
| Unknown | 5,337 | 2,361 | 631 | 1,457 | 1,171 | 638 | 100 | 292 | 2,371 | 969 | 88 | 197 |
| **Female** | 2,692 (49%) | 1,805 (68%) | 571 (82%) | 1,403 (81%) | 786 (64%) | 526 (68%) | 88 (79%) | 294 (81%) | 1,603 (66%) | 871 (73%) | 86 (76%) | 228 (84%) |

**Other:** Other relative; Friend, neighbor, or someone known through family, friends, work, or community; Paid caregiver, health care provider, or clinician; Other

**†** Patient and co-participant age at visit when patients were first diagnosed with MCI or dementia, or last visit if cognitively unimpaired.

**Supplementary Table 3: Association of risk indicator burden (continuous) with incident dementia**

|  | Cox Proportional Hazard Model* | | | | Fine and Gray Model | | | |
| --- | --- | --- | --- | --- | --- | --- | --- | --- |
|  | β | SE | Statistic | P | β | SE | Statistic | P |
| Risk Indicator Burden | 0.29 | 0.04 | 7.27 | 3.57E-13 | 0.31 | 0.05 | 6.76 | 1.4E-11 |
| Cohort |  |  |  |  |  |  |  |  |
| NACC | - | - | - | - | - | - | - | - |
| ADNI | -0.18 | 0.08 | -2.16 | 3e-02 | -0.18 | 0.11 | -1.56 | 0.12 |
| Race/Ethnicity |  |  |  |  |  |  |  |  |
| NLW | - | - | - | - | - | - | - | - |
| Asian | -0.84 | 0.71 | -1.18 | 2e-01 | -0.76 | 0.63 | -1.2 | 0.23 |
| Black | -0.39 | 0.13 | -2.92 | 4e-03 | -0.4 | 0.13 | -2.99 | 0.0028 |
| Latinx | -0.3 | 0.17 | -1.78 | 8e-02 | -0.25 | 0.16 | -1.6 | 0.11 |
| Other | -0.42 | 0.25 | -1.7 | 9e-02 | -0.4 | 0.24 | -1.69 | 0.09 |
| CDR-SB | 0.98 | 0.03 | 38.26 | <0.001 | 0.97 | 0.05 | 18.59 | <0.001 |
| *c-index (se): 0.86 (0.006)  Hazard ratios estimated by exponentiating the beta coefficient from the Cox model (HR = exp(β)) | | | | | | | | |

**Supplementary Table 4: Association of risk indicator burden (categorical) with incident dementia**

|  | Cox Proportional Hazard Model* | | | | Fine and Gray Model | | | |
| --- | --- | --- | --- | --- | --- | --- | --- | --- |
|  | β | SE | Statistic | P | β | SE | Statistic | P |
| Risk Indicator Burden | |  |  |  |  |  |  |  |
| 0 | - | - | - | - | - | - | - | - |
| 1 | 0.24 | 0.13 | 1.88 | 0.06 | 0.28 | 0.14 | 2.04 | 0.041 |
| 2 | 0.6 | 0.13 | 4.76 | 2.0e-06 | 0.64 | 0.14 | 4.69 | 2.7E-06 |
| 3 | 0.72 | 0.15 | 4.82 | 1.4e-06 | 0.78 | 0.17 | 4.57 | 4.8E-06 |
| 4 | 1.61 | 0.26 | 6.25 | 4.1e-10 | 1.65 | 0.24 | 7 | 2.5E-12 |
| Cohort |  |  |  |  |  |  |  |  |
| NACC | - | - | - | - |  |  |  |  |
| ADNI | -0.18 | 0.08 | -2.22 | 0.03 | -0.18 | 0.11 | -1.67 | 0.096 |
| Race/Ethnicity | |  |  |  |  |  |  |  |
| NLW | - | - | - | - | - | - | - | - |
| Asian | -0.84 | 0.71 | -1.19 | 0.23 | -0.76 | 0.64 | -1.19 | 0.23 |
| Black | -0.39 | 0.13 | -2.9 | 3.7e-03 | -0.4 | 0.13 | -3 | 0.0027 |
| Latinx | -0.3 | 0.17 | -1.79 | 0.07 | -0.25 | 0.16 | -1.62 | 0.11 |
| Other | -0.42 | 0.25 | -1.69 | 0.09 | -0.4 | 0.24 | -1.7 | 0.089 |
| CDR-SB | 0.99 | 0.03 | 38.12 | 7.0e-318 | 0.98 | 0.05 | 19.46 | 0 |
| *c-index (se): 0.86 (0.006)  Hazard ratios estimated by exponentiating the beta coefficient from the Cox model (HR = exp(β)) | | | | | | | | |

**Supplementary Figures**

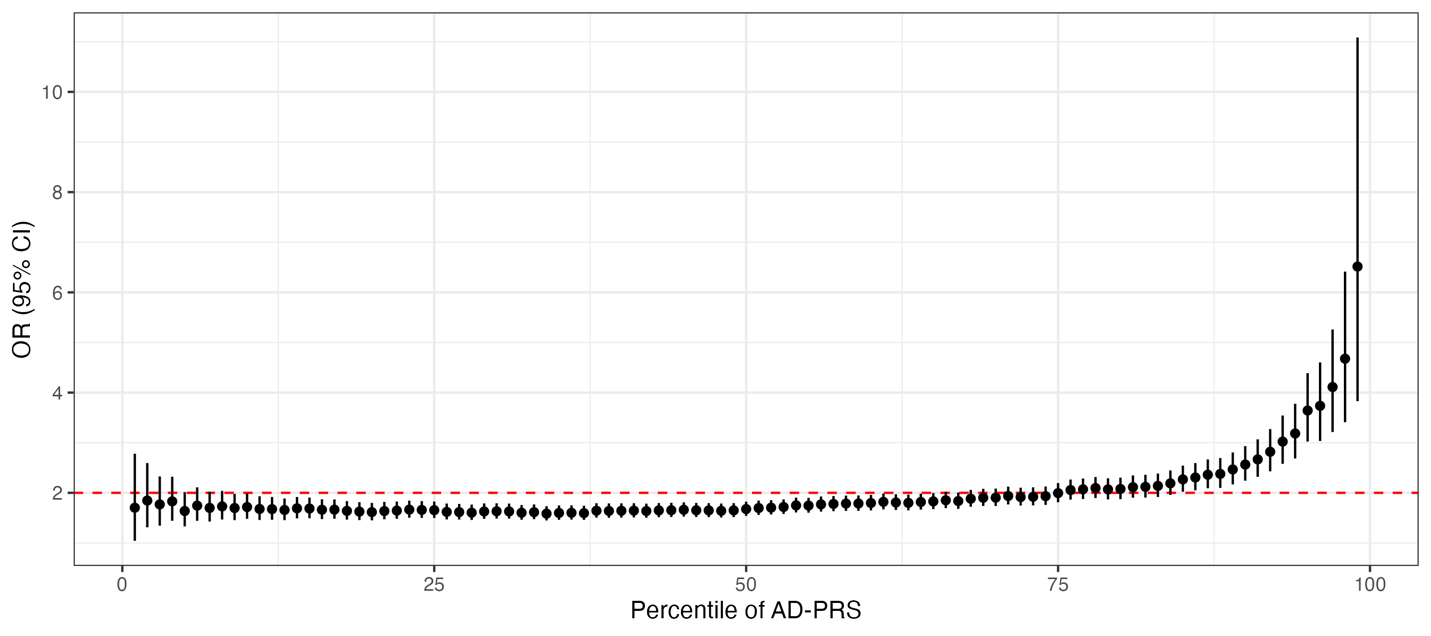

**Supplementary Figure 1:** Logistic regression coefficients for the AD-PRS across percentiles. At the 85^th^ percentile, the high-risk category is associated with increased dementia of OR (95% CI) = 2.27 (2.02, 2.54), p = 1.13e-44. The AD-PRS was constructed in the ADSP using data from 24 cohorts, excluding NACC and ADNI, comprising 11,899 participants (30% cases; mean age = 77 ± 7 years; 62% female; 18% AFR, 42% AMR, 26% EUR, and 14% SAS ancestry).
